# Performance of prediction models for short term outcome in COVID-19 patients in the emergency department: a retrospective study

**DOI:** 10.1101/2020.11.25.20238527

**Authors:** Paul M.E.L. van Dam, Noortje Zelis, Sander M.J. van Kuijk, Aimée E.M.J.H. Linkens, Renée R.A.G. Bruggemann, Bart Spaetgens, Iwan C.C. van der Horst, Patricia M. Stassen

## Abstract

**Introduction:** Coronavirus disease 2019 (COVID-19) has a high burden on the healthcare system and demands information on the outcome early after admission to the emergency department (ED). Previously developed prediction models may assist in triaging patients when allocating healthcare resources. We aimed to assess the value of several prediction models when applied to COVID-19 patients in the ED.

**Methods:** All consecutive COVID-19 patients who visited the ED of a combined secondary/tertiary care center were included. Prediction models were selected based on their feasibility. The primary outcome was 30-day mortality, secondary outcomes were 14-day mortality, and a composite outcome of 30-day mortality and admission to the medium care unit (MCU) or the intensive care unit (ICU). The discriminatory performance of the prediction models was assessed using an area under the receiver operating characteristic curve (AUC).

**Results:** A total of 403 ED patients were diagnosed with COVID-19. Within 30 days, 95 patients died (23.6%), 14-day mortality was 19.1%. Forty-eight patients (11.9%) were admitted to the MCU, 66 patients (16.4%) to the ICU and 152 patients (37.7%) met the composite endpoint. Eleven models were included: RISE UP score, 4C mortality score, CURB-65, MEWS, REMS, abbMEDS, SOFA, APACHE II, CALL score, ACP index and Host risk factor score. The RISE UP score and 4C mortality score showed a very good discriminatory performance for 30-day mortality (AUC 0.83 and 0.84 respectively, 95% CI 0.79-0.88 for both), for 14-day mortality (AUC 0.83, 95% CI: 0.79-0.88, for both) and for the composite outcome (AUC 0.79 and 0.77 respectively, 95% CI 0.75-0.84). The discriminatory performance of the RISE UP score and 4C mortality score was significantly higher compared to that of the other models.

**Conclusion:** The RISE UP score and 4C mortality score have good discriminatory performance in predicting adverse outcome in ED patients with COVID-19. These prediction models can be used to recognize patients at high risk for short-term poor outcome and may assist in guiding clinical decision-making and allocating healthcare resources.

## Background

To mitigate the burden on the healthcare system caused by the Coronavirus disease 2019 (COVID-19) pandemic, it is necessary to identify patients who are at high risk of poor outcomes early in the course of the disease.^1-3^ Although most patients with COVID-19 develop only mild symptoms, some develop severe and potentially fatal complications.^1,2,4,5^ Prediction models could help to forecast outcome when patients present to the emergency department (ED) and may assist in triaging patients when allocating healthcare resources.

Several triage and prediction models have been developed to identify patients with a high risk of adverse outcome.^6-10^ Some of these models were specifically designed for patients with pneumonia (CURB-65) and sepsis (abbreviated Mortality Emergency Department Sepsis (abbMEDS) and sepsis-related organ failure assessment (SOFA)) or for older patients (Risk Stratification in the Emergency Department in Acutely Ill Older Patients (RISE UP)).^6-9^ These models may be useful in patients with COVID-19 as well, as they often present with pneumonia and sepsis and most are older than 65 years. A recent systematic review reported on several new prognostic models specifically designed for patients with COVID-19.^11^ Some models were found to have a reasonable discriminatory performance with an area under the receiver operating characteristic (ROC) curve (AUC) of 0.84.

The present retrospective study aims to validate several previously developed prediction models in patients with COVID-19 in the ED.^6-14^

## Materials and methods

### Study design and setting

This retrospective cohort study was performed at the ED of the Maastricht University Medical Center + (MUMC+). This is a combined secondary/tertiary care center in the Netherlands, with 22,000 ED visits every year. The medical ethics committee of the MUMC+ approved this study (METC 2020-1572). Informed consent was obtained from all individual participants. This study was conducted and reported in line with the Strengthening the Reporting of Observational studies in Epidemiology (STROBE) guidelines.^15^

### Study sample

The study sample consisted of consecutive adult (18 years or older) medical ED patients diagnosed with COVID-19 in the period from March 11^th^ until May 8^th^ 2020. Patients were included if they met the following criteria: 1) symptoms compatible with COVID-19 (i.e., coughing, common cold, sore throat, dyspnea, acute diarrhea, vomiting, fever, or an unexpectedly discovered oxygen saturation below 92%); and 2) positive result of the polymerase chain reaction (PCR) for SARS-CoV-2 in respiratory specimens, or 3) (very) high suspicion of COVID-19 according to the chest computed tomography (CT) scan (CO-RADS 4 or CO-RADS 5).^16^ We excluded patients who revisited the ED after an earlier ED presentation during the study period.

### Data collection

From electronic medical records, we collected data on age, sex and information regarding comorbidity according to the Charlson Comorbidity Index (CCI).^17^ We also retrieved the following vital signs: heart rate (HR), systolic blood pressure (SBP), mean arterial blood pressure (MAP), respiratory rate (RR), oxygen saturation, temperature, and Glasgow Coma Scale (GCS). The Alert Verbal Pain Unresponsive (AVPU) scale was derived from the GCS.^18^ If RR or GCS were missing, we used paCO_2_ and descriptions in the medical records to deduce these values, similar to other studies.^6,14,19^ In addition, we collected routinely assessed laboratory tests: hemoglobin, hematocrit, leukocytes, thrombocytes, lymphocytes, D-dimer, blood gas analysis, bicarbonate, sodium, potassium, blood urea nitrogen (BUN), creatinine, lactate dehydrogenase (LDH), bilirubin, albumin and C -reactive protein (CRP). If hematocrit and pO2 values were missing, we used hemoglobin and oxygen saturation to calculate these values, similar to other studies.^20,21^

Furthermore, we collected the results of the PCR for SARS-CoV-2 in respiratory specimens and the results of the chest CT scan.^16^ Finally, we retrieved data on length of hospital stay, admission to the medium care unit (MCU) or intensive care unit (ICU), and 30-day and 14-day mortality. Data on mortality were verified using the medical records, which are connected to the municipal administration office.

### Prediction models

We searched PubMed for studies on prediction models focusing on patients with COVID-19 using a combination of methodological search terms (prognostic, prediction model, score, regression) and COVID-19 search terms (COVID-19, SARS-CoV-2, coronavirus). In addition, we checked reference lists of manuscripts we identified this way. The search was performed on June 17^th^ and repeated on September 11^th^ to check for more recent publications.

We selected prediction models based on the inclusion of variables that are readily available in the ED and the aim to predict the risk of mortality or progression to severe illness (i.e., tachypnea, hypoxia, and ICU admission with shock, mechanical ventilation, or organ failure). We excluded models that were not clearly described or were not feasible in our ED setting. Prediction models were also excluded if the included variables or the risk calculation were unclear. Models developed using machine learning techniques other than regression and radiologic models were excluded, because these could not be reproduced in our setting.

### Outcomes

The primary outcome was all-cause mortality within 30 days of ED presentation. The secondary outcomes were all-cause mortality within 14 days, and a composite outcome of 30-day mortality and admission to the MCU/ICU. In our hospital, all patients admitted to the ICU were mechanically ventilated.

### Statistical analysis

Baseline characteristics were analyzed using descriptive statistics on the observed data. For each patient, we completed variables of the included prediction models. When the score could not be completed in over 5% of patients due to missing values, data were imputed using stochastic regression imputation. We calculated the AUC under the ROC curve to quantify the discriminatory performance of the included prediction models. An AUC of 0.5 corresponds with very poor discriminatory performance, whereas an AUC of 1.0 means perfect accuracy. We compared the AUCs of the included models using the method of DeLong. All data were analyzed using IBM SPPS Statistics for Windows, IBM Corporation, Armonk N.Y., USA, version 25.0.

## Results

### Study sample

During the study period, 415 ED patients met the inclusion criteria. After the exclusion of twelve patients because of refusal of informed consent, we included 403 patients for analysis (Table 1). The median age of patients was 71 years (IQR 60-78) and 255 patients (63.2%) were older than 65 years. Most patients (66.0%) were male. The PCR for SARS-CoV-2 was positive in 323 patients (80.1%) and the chest CT scan was positive in 325 patients (80.6%). A total of 307 patients (76.2%) were admitted to the hospital, whereas the other patients were discharged home for further recovery. The median length of hospital stay was 6 days (IQR 3-12).

**Table 1.** Characteristics of the study sample.

|  | Reference values | Study sample (n = 406) |
| --- | --- | --- |
| Age, median (IQR), years |  | 71 (60-78) |
| Male, n% |  | 266 (66.0) |
| Symptoms upon ED visit, n% |  |  |
| Fever |  | 249 (61.8) |
| Cough |  | 287 (71.2) |
| Dyspnea |  | 278 (69.0) |
| General malaise |  | 187 (46.4) |
| Headache |  | 52 (12.9) |
| Thoracic pain |  | 61 (15.1) |
| Nausea |  | 92 (22.8) |
| Diarrhea |  | 90 (22.3) |
| No respiratory symptoms |  | 55 (13.6) |
| PCR positive, n% |  | 323 (80.1) |
| Chest CT scan positive, n% |  | 325 (80.6) |
| Outcomes |  |  |
| Admission to hospital, n% |  | 307 (76.2) |
| Length of hospital stay, median (IQR), days |  | 6 (3-12) |
| Treatment restrictions, n% |  | 142 (35.2) |
| Mortality within 14 days, n% |  | 77 (19.1) |
| Mortality within 30 days, n% |  | 95 (23.6) |
| MCU admission, n% |  | 48 (11.9) |
| ICU admission, n% |  | 66 (16.4) |
| Composite endpoint (mortality and/or admission to MCU/ICU), n% |  | 152 (37.7) |

CT, computed tomography; ED, emergency department; ICU, intensive care unit; IQR, interquartile range; MCU, medium care unit; PCR, polymerase chain reaction; SD, standard deviation.

In our sample, 66 patients (16.4%) were admitted to the ICU, 48 patients (11.9%) to the MCU, and 95 patients died during follow up, yielding a 30-day mortality of 23.6% and a 14-day mortality of 19.1%. The survival curve is shown in Figure 1. A total of 152 patients (37.7%) met the composite endpoint of 30-day mortality and admission to MCU/ICU.

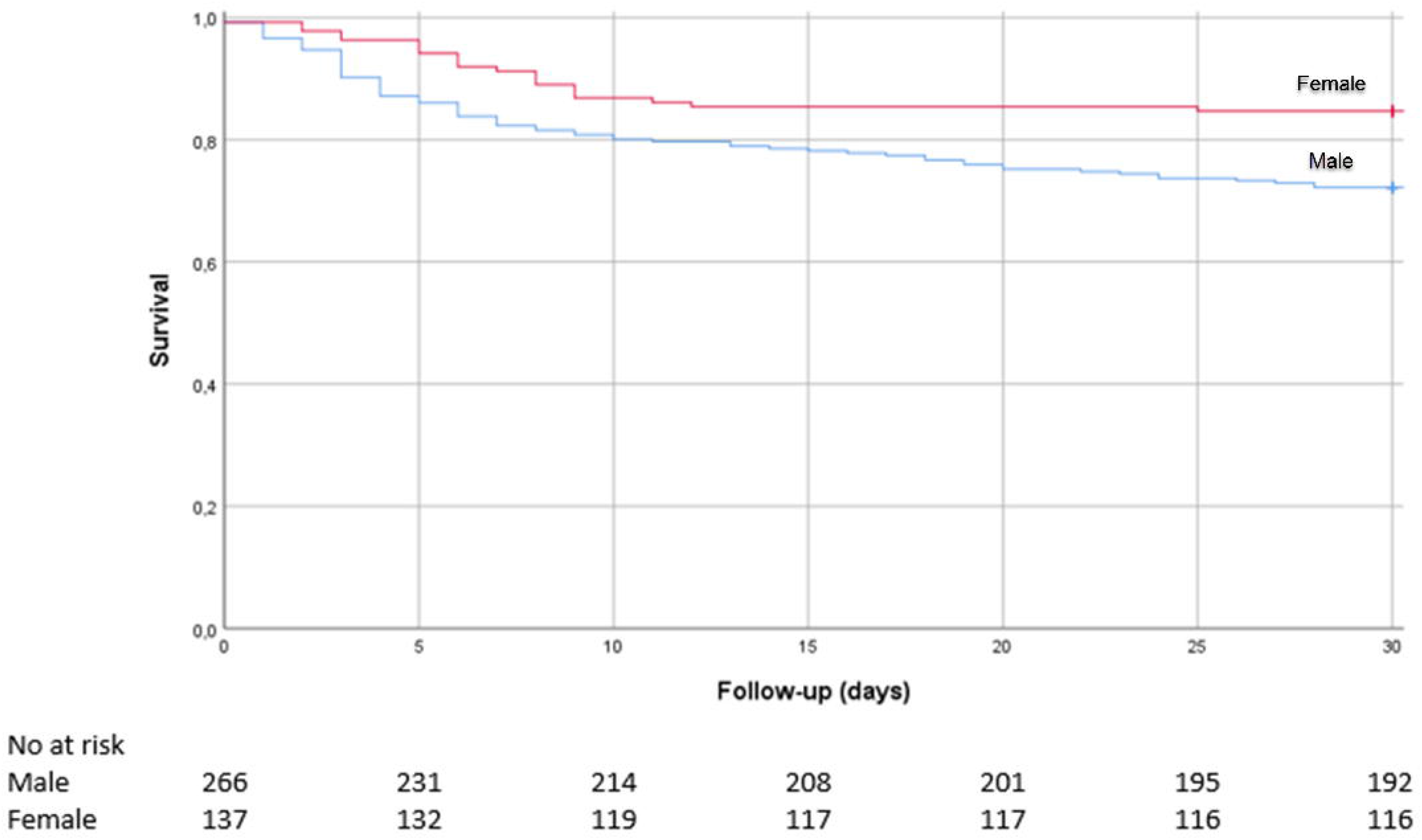

### Prediction models

We included eleven prediction models (Table 2), of which seven prediction models were not explicitly developed for patients with COVID-19: RISE UP, CURB-65, Modified Early Warning Score (MEWS), Rapid Emergency Medicine Score (REMS), abbMEDS, SOFA and Acute Physiology And Chronic Health Evaluation II (APACHE II).^6-10,12-14^ Furthermore, in a recent systematic review, 16 prediction models specifically designed for patients with COVID-19 were identified.^11^ Of these models, eight estimated mortality risk in patients with suspected or confirmed COVID-19, five aimed to predict progression to severe disease and three estimated length of hospital stay. We excluded 14 of these models for the following reasons: no clear description of the variables or risk calculation (n = 5), not compatible with our setting because of the use of machine learning (n = 5), or inclusion of radiologic characteristics (n = 4). We included two prognostic models from the systematic review (ACP score and Host risk factor score).^22,23^ Additionally, we included two more recently published prediction models (CALL score and the Coronavirus Clinical Characterisation Consortium (4C) mortality score) not included in the systematic review.^24,25^

**Table 2.** Overview of included prediction models.

| Prediction model | Original population | Items | Outcome |
| --- | --- | --- | --- |
| RISE UP<sup>6</sup> | Older patients (>65 years) | Age, HR, MAP, RR, O<sub>2</sub> saturation, GCS, BUN, bilirubin, albumin, LDH | Mortality (30-day) |
| 4C mortality score<sup>25</sup> | Patients with COVID-19 | Age, sex, comorbidity, RR, GCS, O<sub>2</sub> saturation, BUN, CRP | Mortality (in hospital) |
| CURB-65<sup>7</sup> | Patients with pneumonia | Age, confusion, BUN, RR, blood pressure | Mortality (30-day) |
| MEWS<sup>10</sup> | Patients in the ED | HR, blood pressure, O<sub>2</sub> saturation, RR, temperature, AVPU | Mortality and ICU admission (<sup>a</sup>) |
| REMS<sup>14</sup> | Patients in the ED | Age, MAP, HR, RR, GCS, O<sub>2</sub> saturation | Mortality (<sup>a</sup>) |
| abbMEDS<sup>9</sup> | Sepsis patients | Age, comorbidity, confusion, septic shock, thrombocytes, pneumonia, respiratory symptoms, confusion, nursing home | Mortality (28-day) |
| SOFA<sup>12</sup> | ICU patients | pO<sub>2</sub>/FiO<sub>2</sub> ratio, GCS, MAP, thrombocytes, bilirubin, creatinine | Mortality (<sup>a</sup>) |
| APACHE II<sup>13</sup> | ICU patients | Age, HR, RR, MAP, GCS, temperature, hematocrit, pH, pO<sub>2</sub>, leucocytes, sodium, potassium, creatinine, comorbidity | Mortality (<sup>a</sup>) |
| CALL score<sup>24</sup> | Patients with COVID-19 | Age, comorbidity, lymphocytes, LDH | Progression to severe disease (<sup>a, b</sup>) |
| ACP index<sup>22</sup> | Patients with COVID-19 | Age, CRP | Mortality (12-day) |
| Host risk factor score<sup>23</sup> | Patients with COVID-19 | Age, sex, hypertension | Progression to severe disease (<sup>a, b</sup>) |

4C, Coronavirus Clinical Characterisation Consortium; abbMEDS, abbreviated Mortality Emergency Department Sepsis; ACP, Age C-reactive Protein; APACHE II, Acute Physiology and Chronic Health Evaluation II; AVPU, Alert Verbal Pain Unresponsive; BUN, blood urea nitrogen; CALL, Comorbidity Age Lymphocyte LDH; CRP, C-reactive protein; CURB-65, Confusion Urea Respiration Blood pressure; ED, emergency department; GCS, Glasgow coma scale; HR, heart rate; ICU, intensive care unit; LDH, lactate dehydrogenase; MAP, mean arterial pressure; MEWS, Modified Early Warning Score; REMS, Rapid Emergency Medicine Score; RISE UP, Risk Stratification in the Emergency Department in Acutely Ill Older Patients; RR, respiratory rate; SOFA, Sepsis-related Organ Failure Assessment.
<sup>a</sup> Time span not specified in the original article.
<sup>b</sup> Progression to severe disease was not specified in the original article.

A total of six prediction models (RISE UP, 4C mortality, CURB-65, SOFA, APACHE II, and CALL) could not be calculated in >5% of the patients because of missing values (vital signs and laboratory tests). Therefore, missing data were imputed using stochastic regression imputation.

### Validation of the prediction models

The prediction models were used to calculate the risk of an adverse outcome (Table 3, Figure 2). The RISE UP score and 4C mortality score showed the best discriminatory performance and respectively yielded an AUC of 0.83 (95% CI: 0.79-0.88) and 0.84 (95% CI: 0.79-0.88) for 30-day mortality, an AUC of 0.83 (95% CI: 0.79-0.88) and 0.83 (95% CI: 0.79-0.88) for 14-day mortality, and an AUC of 0.79 (95% CI: 0.74-0.84) and 0.77 (95% CI: 0.72-0.82) for the composite endpoint.

**Table 3.** Comparison of the AUCs of included prediction models.

| Prediction model | 30-day mortality (AUC, 95% CI) | Difference with RISE UP (p value)<sup>a</sup> | 14-day mortality (AUC, 95% CI) | Difference with RISE UP (p value)<sup>a</sup> | Composite endpoint (mortality and/or MCU/ICU admission) (AUC, 95% CI) | Difference with RISE UP (p value)<sup>a</sup> |
| --- | --- | --- | --- | --- | --- | --- |
| RISE UP | 0.83 (0.79-0.88) | reference | 0.83 (0.79-0.88) | reference | 0.79 (0.75-0.84) | reference |
| 4C mortality score | 0.84 (0.79-0.88) | 0.914 | 0.83 (0.79-0.88) | 0.926 | 0.77 (0.72-0.82) | 0.170 |
| CURB-65 | 0.75 (0.70-0.80) | <0.001 | 0.75 (0.70-0.81) | 0.005 | 0.68 (0.62-0.73) | <0.001 |
| MEWS | 0.64 (0.58-0.70) | <0.001 | 0.62 (0.56-0.69) | <0.001 | 0.73 (0.68-0.78) | 0.037 |
| REMS | 0.73 (0.68-0.78) | <0.001 | 0.74 (0.69-0.79) | 0.001 | 0.72 (0.66-0.77) | <0.001 |
| abbMEDS | 0.75 (0.70-0.81) | <0.001 | 0.75 (0.69-0.81) | 0.003 | 0.71 (0.66-0.76) | <0.001 |
| SOFA | 0.72 (0.67-0.78) | <0.001 | 0.72 (0.65-0.78) | <0.001 | 0.76 (0.71-0.81) | 0.200 |
| APACHE II | 0.71 (0.65-0.78) | <0.001 | 0.73 (0.67-0.79) | 0.004 | 0.69 (0.63-0.74) | <0.001 |
| CALL score | 0.76 (0.71-0.81) | 0.002 | 0.76 (0.70-0.81) | 0.012 | 0.70 (0.65-0.75) | <0.001 |
| ACP index | 0.67 (0.61-0.73) | <0.001 | 0.67 (0.61-0.73) | <0.001 | 0.66 (0.61-0.72) | <0.001 |
| Host risk factor score | 0.64 (0.57-0.70) | <0.001 | 0.62 (0.56-0.69) | <0.001 | 0.63 (0.57-0.68) | <0.001 |

4C, Coronavirus Clinical Characterisation Consortium; abbMEDS, abbreviated Mortality Emergency Department Sepsis; ACP, Age C-reactive protein; APACHE II, Acute Physiology and Chronic Health Evaluation II; AUC, area under the curve; CALL, Comorbidity Age Lymphocyte LDH; CI, confidence interval; CURB-65, Confusion Urea Respiration Blood pressure; ICU, intensive care unit; MCU, medium care unit; MEWS, Modified Early Warning Score; REMS, Rapid Emergency Medicine Score; RISE UP, Risk Stratification in the Emergency Department in Acutely Ill Older Patients; SOFA, Sepsis-related Organ Failure Assessment.
<sup>a</sup> Comparison of the AUC of the prediction model with the AUC of the RISE UP score.

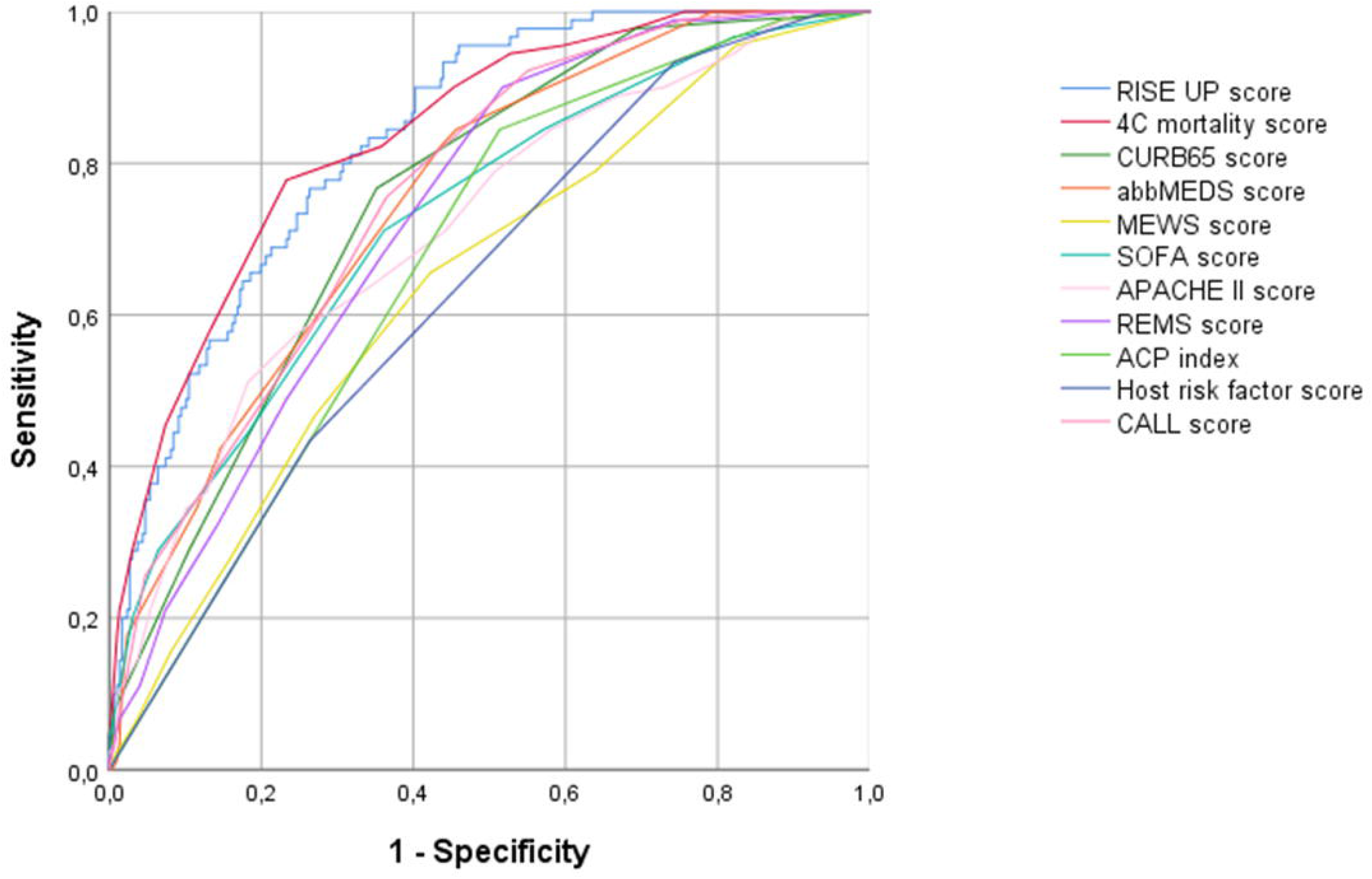

In comparison, the CURB-65, MEWS, REMS, abbMEDS, SOFA, APACHE II, CALL, ACP and Host risk factor score yielded AUCs ranging from 0.64 to 0.76 for 30-day mortality, AUCs ranging from 0.62 to 0.76 for 14-day mortality, and AUCs ranging from 0.68 to 0.76 for the composite endpoint. The discriminatory performance of the RISE UP score and 4C mortality score was significantly higher than that of the other models using the DeLong method.

## Discussion

In this retrospective study, we externally validated eleven prediction models for their ability to predict 30-day mortality or admission to MCU/ICU in ED patients with COVID-19. We found that both the RISE UP and 4C mortality score had very good discriminatory performance, which was the highest of the models we analyzed. The model yielded high AUCs for both 14-day mortality (both AUC of 0.83) and 30-day mortality (AUC of 0.83 and 0.84). The nine other models showed significantly lower discriminatory performance. The CURB-65, REMS, abbMEDS, SOFA, APACHE II, and CALL score had a good discriminatory performance (AUC ranging from 0.71 to 0.76) as well, while the ACP index and Host risk factor score had a moderate to poor performance (AUC of 0.67 and 0.64, respectively). Most prediction models had a higher discriminatory performance for predicting mortality than for predicting the composite outcome of mortality and MCU/ICU admission.

The RISE UP score was recently developed to predict 30-day all-cause mortality in older medical ED patients and consists of easily and readily available items during the ED visit.^6^ It is not unexpected that the model works well for admitted patients with COVID-19, since many of these patients in our cohort (63.2%) were 65 years or older. High mortality in older patients with COVID-19 was shown previously.^26-29^ The 4C mortality score was recently developed to predict in-hospital mortality in a very large cohort of COVID-19 patients in the UK.^25^ The good discriminatory performance of both the RISE UP and 4C mortality scores can be explained because these models include items that reflect the severity of illness in ED patients and are indicative of sepsis, organ failure and/or shock (i.e. abnormal vital signs, LDH, BUN, Bilirubin). The items of the RISE UP and 4C mortality score are quite similar. Elevated levels of LDH were found to predict adverse outcomes in patients with COVID-19.^30^ The prognosis of ED patients is reflected by the presentation of the patients at the ED, which results from both the severity of the current disease and preexisting factors (i.e. age and comorbidities).^1,4^ Regarding feasibility, the probability of a poor outcome can be predicted in the first two hours of the ED visit by both models. One disadvantage of the 4C mortality score may be that it contains the number of comorbidities of the ED patients, which is not always available in the ED. This is a disadvantage compared to the RISE UP score, which consists of six items that are readily available in the ED. Moreover, the RISE UP score can easily be implemented with an online calculator (https://jscalc.io/calc/o1vzp36bIDGQUCYl). To guide clinical decision-making, prediction models that can be computed easily and quickly are of great value.

The CURB-65 is commonly used to assess the severity and mortality in patients with community-acquired pneumonia.^7^ In our cohort, we found that the score had a moderate to good ability to discriminate between mortality and survival (AUC of 0.75). In other studies in patients with COVID-19, the CURB-65 score was found to have very good discriminatory performance for mortality and progression to severe disease with AUCs ranging from 0.81 to 0.88.^31-34^ The highest AUC (0.88) was found in a Turkish study.^32^ Their high AUC may be explained by the inclusion of patients with less severe COVID-19 (more often lower CURB scores and lower mortality) compared to our patients. The MEWS and REMS were designed for early detection of high-risk patients by assigning points to vital signs and can both be easily applied in the ED. In our cohort, the MEWS score showed only reasonable discriminatory performance (AUC of 0.64), while the REMS score yielded moderate to good performance (AUC of 0.73). In one Chinese study, the MEWS score and REMS score were analyzed in 138 patients with COVID-19.^35^ The MEWS showed an AUC of 0.68, similar to the AUC in our sample. The REMS score was found to have an AUC of 0.84. Our patients were older than the patients in the Chinese study (median 71 versus 58 years), which probably explains the higher AUC, as the AUC was 0.77 in the 50 Chinese patients older than 65 years.

APACHE II and SOFA scores are used to predict mortality in ICU patients. The discriminatory performance of these scores in our cohort was moderate to good (AUC of 0.71 and 0.72, respectively). These findings were comparable to those reported in other studies with patients with COVID-19.^30,31,36^ In one Chinese study in ICU patients with COVID-19, the AUC of the APACHE II score was 0.97 and the AUC of the SOFA score was 0.87, which is much higher than the AUCs we found.^31^ However, our patients were less frequently admitted the ICU (only 16.4%). Consequently, our population is more heterogeneous and mortality is probably more difficult to predict. The APACHE II score turned out to be less feasible in an ED setting because in our ED, an arterial blood gas is measured on indication only (in 37.5% of our patients no arterial blood gas was measured).

The three other prognostic models that were specifically designed for patients with COVID-19 had varying predictive performances in our cohort. The CALL score had good predictive value (AUC of 0.76). This CALL score was developed to predict progression to severe disease in the first 5 to 10 days in a cohort of 208 Chinese patients with COVID-19.^24^ The AUC in the Chinese study was 0.86, which was higher than the AUC we found. Application of a new model in an independent cohort usually results in a lower AUC, and in addition, the patients in the Chinese cohort were much younger than our patients (mean 44 versus 71 years) and their follow-up period was shorter. The ACP index was developed to predict 12-day mortality in patients with COVID-19 in Wuhan.^22^ The Host risk factor score was developed to predict mortality or progression to severe disease.^23^ The discriminatory performance of these two scores was not reported by the authors. In our external validation, both scores had poor discriminatory performance (AUC of 0.67 (ACP index) and 0.64 (Host risk factor score)). In a recent Spanish study in nursing home residents, the ACP and Host risk factor score yielded comparable low AUCs (AUC of 0.60 and 0.55, respectively).^34^ The difference between our study and the original Chinese studies may also be explained by the different phase of the COVID-19 pandemic in which the studies took place, as in Europe, physicians were already slightly more prepared and outcomes may therefore differ.

Our study had several limitations. First, our study was performed in a single medical center, which may limit the generalizability of the results. However, our cohort of patients with COVID-19 was relatively large and has been recruited in one of the most heavily affected areas of the Netherlands. Furthermore, by validating all prediction models in the same cohort, there were no differences in the patient sample, and we could truly compare the scores.^37^ Second, the process of selecting prediction models for our analysis might have been incomplete. We chose prediction models that were feasible in our ED setting, which may be different for other EDs. Third, in a subgroup of patients with preexisting frailty or severe comorbidity, it was decided to initiate conservative care only (35.2% had treatment restrictions). As these decisions may be different in other countries, we decided to study MCU/ICU admissions as a composite outcome only. In addition, we decided to calculate an AUC for 14-day and 30-day mortality in the 261 patients without treatment restrictions (Supplementary Table). We found comparable AUCs for both outcomes (AUC of 0.82 and 0.84 for the RISE UP, respectively). We therefore found no evidence for differences in performance of the models between patients with and without treatment restrictions. Last, the number of ICU and MCU admissions in our study was relatively low (16.4% and 11.9%, respectively). In our cohort, 23.8% of the patients were discharged home and therefore not able to reach this endpoint. However, these patients were apparently judged to be not severely ill and 30-day mortality in this subgroup was low (3.1%).

## Conclusion

In conclusion, the RISE UP and 4C mortality score had the highest discriminatory performance for short term mortality in ED patients with COVID-19. Prediction models like the RISE UP and 4C mortality score are useful to identify patients at high risk for adverse outcomes and may be a first step in guiding clinical decision-making and allocating healthcare resources in this pandemic, in which we have to deal with scarcity of clinical facilities and materials. However, this needs to be subject of further investigation.

## Data Availability

All data referred to in the manuscript are available upon reasonable request.

## Contributors

PD, NZ, and PMS collected the clinical data. PD, NZ and SMJK performed the statistical analysis. All authors interpreted data. PD drafted the first version of the manuscript. NZ, IH, SMJK, RB, AL, BS and PMS critically reviewed the manuscript. All authors have read and approved the final version of the manuscript.

---

**Supplementary Table 1.** Comparison of the AUCs of included prediction models in subgroup analysis (patients without treatment restrictions only, n = 261).

| Prediction model | 30-day mortality (AUC, 95% CI) | Difference with RISE UP (p value) <sup>a</sup> | 14-day mortality (AUC, 95% CI) | Difference with RISE UP (p value) <sup>a</sup> | Composite endpoint (mortality and/or MCU/ICU admission) (AUC, 95% CI) | Difference with RISE UP (p value) <sup>a</sup> |
| --- | --- | --- | --- | --- | --- | --- |
| RISE UP | 0.84 (0.76-0.91) | reference | 0.82 (0.72-0.92) | reference | 0.81 (0.75-0.87) | reference |
| 4C Mortality score | 0.85 (0.77-0.94) | 0.617 | 0.84 (0.75-0.93) | 0.644 | 0.77 (0.71-0.84) | 0.148 |
| CURB-65 | 0.75 (0.64-0.86) | 0.039 | 0.75 (0.62-0.87) | 0.156 | 0.65 (0.58-0.73) | <0.001 |
| MEWS | 0.63 (0.49-0.76) | <0.001 | 0.58 (0.40-0.75) | <0.001 | 0.78 (0.71-0.85) | 0.785 |
| REMS | 0.76 (0.67-0.86) | 0.154 | 0.80 (0.73-0.87) | 0.765 | 0.72 (0.65-0.79) | 0.020 |
| abbMEDS | 0.78 (0.69-0.86) | 0.095 | 0.78 (0.68-0.89) | 0.427 | 0.72 (0.65-0.78) | 0.007 |
| SOFA | 0.73 (0.61-0.84) | 0.034 | 0.74 (0.59-0.89) | 0.245 | 0.79 (0.73-0.86) | 0.877 |
| APACHE II | 0.69 (0.56-0.82) | 0.036 | 0.77 (0.64-0.90) | 0.578 | 0.68 (0.61-0.76) | 0.005 |
| CALL score | 0.75 (0.66-0.84) | 0.073 | 0.77 (0.66-0.87) | 0.359 | 0.67 (0.59-0.74) | <0.001 |
| ACP index | 0.73 (0.64-0.82) | 0.008 | 0.73 (0.63-0.83) | 0.053 | 0.68 (0.61-0.75) | <0.001 |
| Host risk factor score | 0.70 (0.60-0.79) | 0.022 | 0.69 (0.57-0.82) | 0.100 | 0.63 (0.59-0.71) | <0.001 |

4C, Coronavirus Clinical Characterisation Consortium; abbMEDS, abbreviated Mortality Emergency Department Sepsis; ACP, Age C-reactive protein; APACHE II, Acute Physiology and Chronic Health Evaluation II; AUC, area under the curve; CALL, Comorbidity Age Lymphocyte LDH; CI, confidence interval; CURB-65, Confusion Urea Respiration Blood pressure; ICU, intensive care unit; MCU, medium care unit; MEWS, Modified Early Warning Score; REMS, Rapid Emergency Medicine Score; RISE UP, Risk Stratification in the Emergency Department in Acutely Ill Older Patients; SOFA, Sepsis-related Organ Failure Assessment.
<sup>a</sup> Comparison of the AUC of the prediction model with the AUC of the RISE UP score.

